# The King’s College London Coronavirus Health and Experiences of Colleagues at King’s Study: SARS-CoV-2 antibody response in a higher education sample

**DOI:** 10.1101/2021.01.26.21249744

**Authors:** Daniel Leightley, Valentina Vitiello, Alice Wickersham, Katrina A.S. Davis, Gabriella Bergin-Cartwright, Grace Lavelle, Sharon A.M Stevelink, Matthew Hotopf, Reza Razavi, On behalf of the KCL CHECK research team

## Abstract

**Objective:** To assess the feasibility of home antibody testing as part of large-scale study, the King’s College London Coronavirus Health and Experiences of Colleagues at King’s (KCL CHECK).

**Methods:** Participants of the KCL CHECK study were sent a SureScreen Diagnostics COVID-19 IgG/IgM Rapid Test Cassette to complete at home in June 2020 (phase 1) and September 2020 (phase 2). Participants were asked to upload a test result image to a study website. Test result images and sociodemographic information were analysed by the research team.

**Results:** A total of n=2716 participants enrolled in the KCL CHECK study, with n=2003 (73.7%) and n=1825 (69.3%) consenting and responding to phase 1 and 2. Of these, n=1882 (93.9%; phase 1) and n=1675 (91.8%; phase 2) returned a valid result. n=123 (6.5%; phase 1) and n=91 (5.4%; phase 2) tested positive for SARS-CoV-2 antibodies. A total of n=1488 participants provided a result in both phases, with n=57 (3.8%) testing positive for SARS-CoV-2 antibodies across both phases, suggesting a reduction in the number of positive antibody results over time. Initial comparisons showed variation by age group, gender and clinical role.

**Conclusions:** Our study highlights the feasibility of rapid, repeated and low-cost SARS-CoV-2 serological testing without the need for face-to-face contact.

**What is already known about this subject?:** Higher education institutions have a duty of care to minimise the spread and transmission of COVID-19 in its campuses, and among staff and students. The reopening of higher education buildings and campuses has brought about a mass movement of students, academics and support staff from across the UK. Serological antibody studies can assist by highlighting groups of people and behaviours associated with high risk of COVID-19.

**What are the new findings?:** We report a framework for SARS-CoV-2 serological antibody testing in an occupational group of postgraduate research students and current members of staff at King’s College London. Over two phases of data collection, 6.5% (phase 1) and 5.4% (phase 2) tested positive for SARS-CoV-2 antibodies, with only 3.8% testing positive for antibodies in both phases, suggesting a reduction in positive antibody results over time.

**How might this impact on policy or clinical practice in the foreseeable future?:** Our study highlights the feasibility of rapidly deploying low-cost and repeatable SARS-CoV-2 serological testing, without the need for face-to-face contact, to support the higher education system of the UK.

## Introduction

In the United Kingdom (UK), a nationwide lockdown was announced on 23 March 2020 in response to the coronavirus pandemic. The lockdown initially lasted for three weeks but has since been extended and modified to account for regional outbreaks. This has raised many questions as to how the higher education (HE) sector in the UK can cope with extended periods of lockdown and handle the return of staff and students to campuses.

In the UK, it is estimated that 2.4 million students are currently studying at 165 HE institutions, supported by half a million academic and support staff [1]. A major concern among public health experts is the mass migration of students across the UK, travel to/from campuses for both staff and students, and the risk of spreading SARS-CoV-2. One possible approach could be to identify those who have previously been infected with SARS-CoV-2 in order to highlight characteristics and behaviours associated with high risk of COVID-19 and consider these factors in measures to reduce the risk of transmission.

Establishing the presence of SARS-CoV-2 antibodies poses significant challenges to the research community, HE employers and healthcare providers in light of social distancing measures which impede in-person testing. Antibody tests can be used to indicate whether a person is likely to have already had SARS-CoV-2. Validation studies have shown that specific IgM and or IgG antibodies can be detectable from 4-5 days post-symptom development (evaluated in a hospital setting), with positive IgM antibodies in 70% of symptomatic patients by day 8-14 and 90% of antibody tests positive by day 11-24 [2].

In this short preliminary communication, we investigate 1) the feasibility of home antibody testing as part of large-scale cohort study (KCL CHECK) and building upon our preliminary investigation (see [3] for further details); 2) the antibody response of participants at two timepoints; and 3) to stratify antibody results by self-reported occupational and sociodemographic.

## Methods

KCL CHECK seeks to explore the psychological, social and physical impact of SARS-CoV-2 in a longitudinal cohort of postgraduate research students and members of staff at King’s College London, a large Russell Group University in London, UK (for study protocol see [4]). Some current members of staff in the cohort also hold joint contracts with the UK National Health Service. Participants were recruited by email in April 2020 and volunteered to complete surveys and antibody testing over the subsequent 18-month study period. While there was no incentivisation, participation could have been motivated by the offer of antibody testing.

A Rapid Immunoglobulin Test Cassette, a lateral flow immunoassay, was used to detect the presence of IgM and IgG antibodies to the ‘spike’protein, thereby providing evidence for previous infection with SARS-CoV-2. A detailed analysis of Rapid Immunoglobulin Test Cassettes has previously been reported [5], [6]. SureScreen Diagnostics COVID-19 IgG/IgM Rapid Test Cassettes, the necessary equipment and detailed instructions were sent to participants’home address. Participants were informed that the test was not licenced for clinical use and was being used for research purposes. A reporting sheet included each participant’s unique identifier, such that when participants submitted a photograph of their test result online via the study website, it could be securely linked to their survey results. Participant photographs were then analysed by the research team and rated according to the following scales: ‘positive’, indicating the presence of IgG/IgM (denoted via pink lines on each item); ‘negative’(denoted via a pink line on the control item); and ‘invalid’(denoted by no lines appearing on any item or blood in the buffer zone). Preliminary results for phase 1 home antibody testing for KCL CHECK participants has previously been reported for this study (see [3] for further detail).

Statistical analysis was performed in STATA 16.0. Descriptive statistics and antibody test results are reported as frequencies and unweighted row percentages. Invalid test results were omitted from the analyses.

## Results

A total of n=2716 participants are enrolled in the KCL CHECK study, with n=2288 (84.2%) and n=2284 (84.1%) consenting to receive a SARS-CoV-2 antibody home testing kit, for phase 1 and phase 2, respectively. For phase 1 testing, test cassettes were posted to participants in June 2020, with n=1882 (93.9%) participants returning valid results. For phase 2 testing, test cassettes were posted to participants in September 2020, with n=1675 (91.8%) participants returning valid results. It is important to note that n=224 (9.8%; phase 1) and n=156 (6.8%; phase 2) participants needed their home testing kits to be resent in July 2020 and October 2020 respectively due to technical issues with the test, an invalid test result, or loss in transit.

Overall, n=123 (6.5%) tests were positive in phase 1, and n=91 (5.4%) were positive in phase 2. Stratifying by sociodemographic and occupational characteristics presented particular variation, for example between staff (phase 1: n=90; 6.4% | phase 2: n=68; 5.1%) and post-graduate students (phase 1: n=25; 7.9% | phase 2: n=18; 6.8%) (Table 1). Similarly, there was some variation in positive rates based on having a clinical role but without COVID-19 patient contact (phase 1: n=111; 6.4% | phase 2: n=82; 5.3%) versus having a clinical role with suspected COVID-19 patient contact (phase 1: n=7; 7.1% | phase 2: n=5; 5.9%).

**Table 1.**
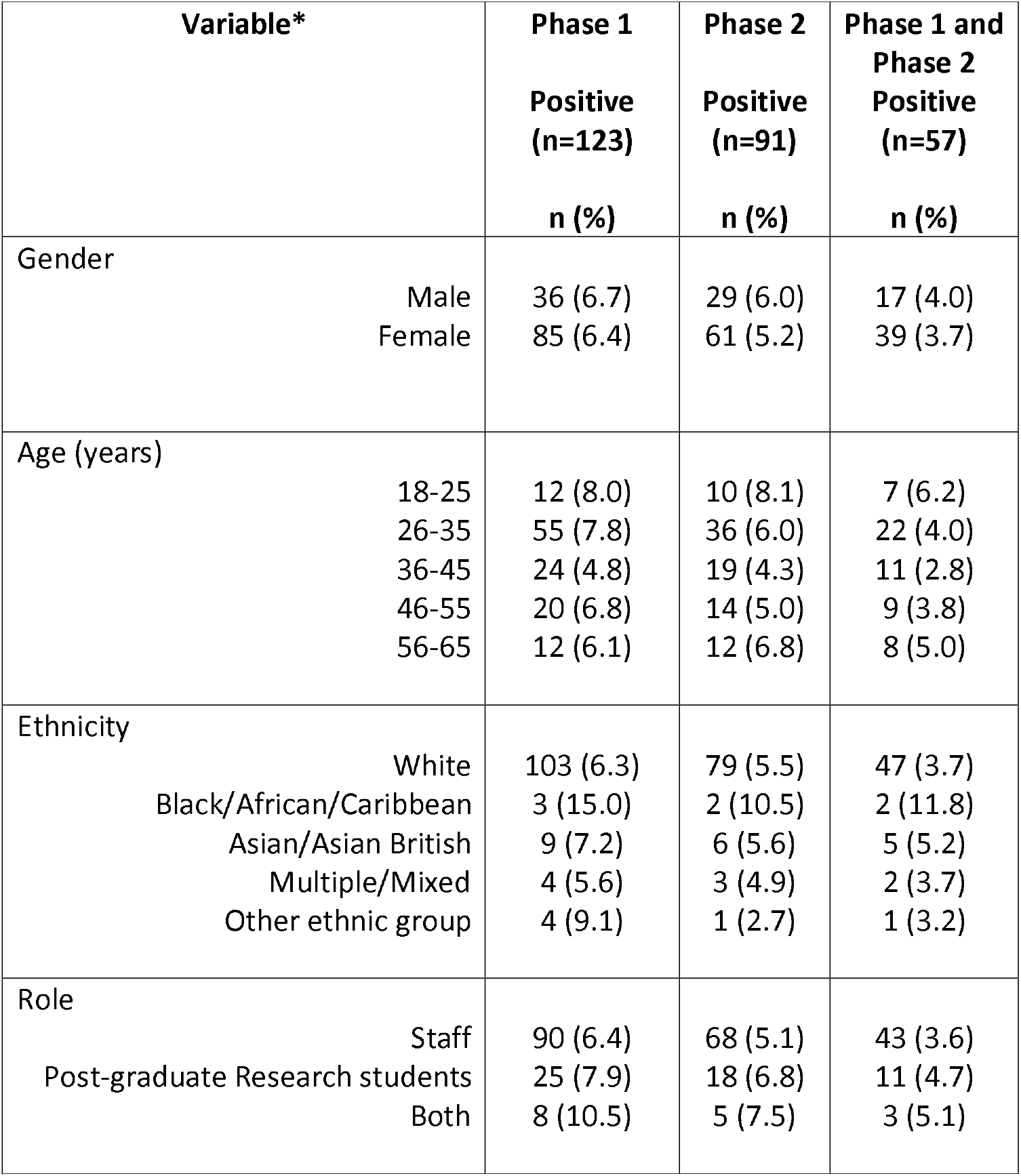
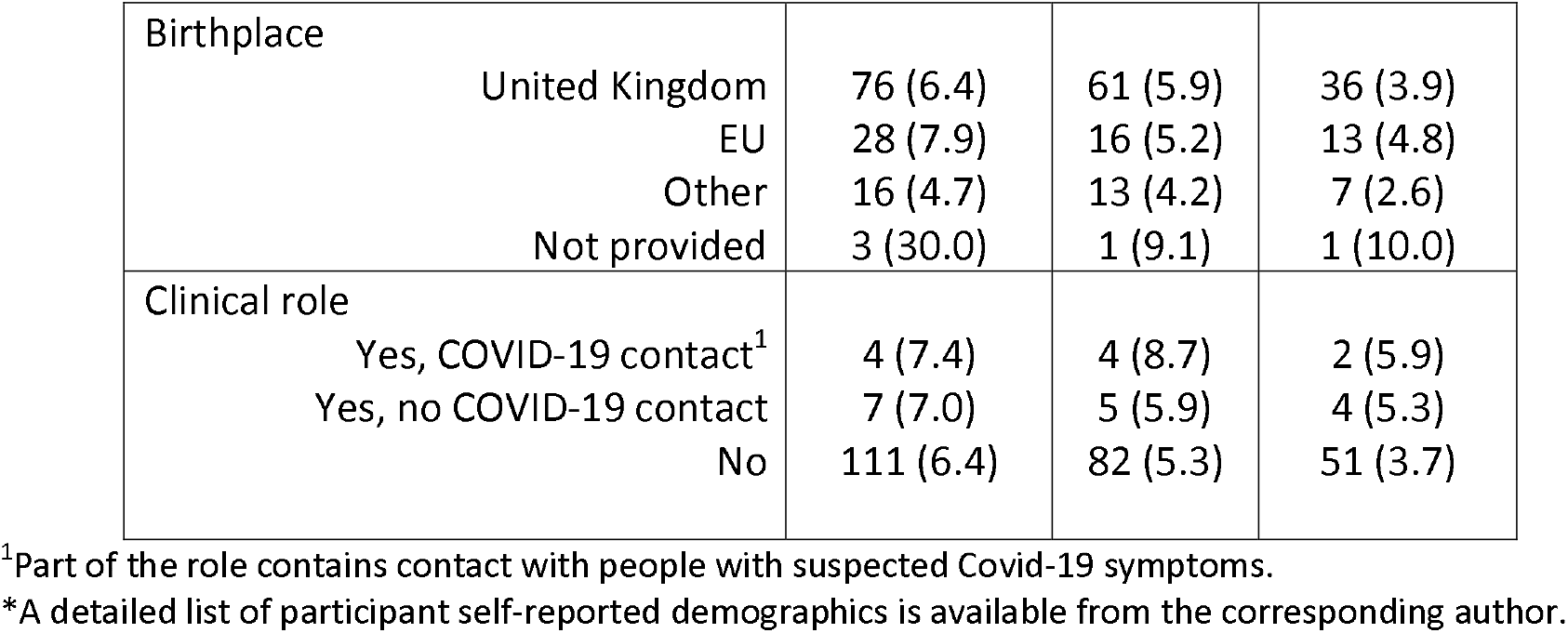
Proportion of SARS-CoV-2 antibody results that were positive, stratified by self-reported socio-demographic and occupational information. Proportion of negative results available from the corresponding author upon request.

| Variable* |  | Phase 1<br>Positive<br>(n=123)<br>n (%) | Phase 2<br>Positive<br>(n=91)<br>n (%) | Phase 1 and<br>Phase 2<br>Positive<br>(n=57)<br>n (%) |
| --- | --- | --- | --- | --- |
| Gender | Male | 36 (6.7) | 29 (6.0) | 17 (4.0) |
|  | Female | 85 (6.4) | 61 (5.2) | 39 (3.7) |
| Age (years) | 18-25 | 12 (8.0) | 10 (8.1) | 7 (6.2) |
|  | 26-35 | 55 (7.8) | 36 (6.0) | 22 (4.0) |
|  | 36-45 | 24 (4.8) | 19 (4.3) | 11 (2.8) |
|  | 46-55 | 20 (6.8) | 14 (5.0) | 9 (3.8) |
|  | 56-65 | 12 (6.1) | 12 (6.8) | 8 (5.0) |
| Ethnicity | White | 103 (6.3) | 79 (5.5) | 47 (3.7) |
|  | Black/African/Caribbean | 3 (15.0) | 2 (10.5) | 2 (11.8) |
|  | Asian/Asian British | 9 (7.2) | 6 (5.6) | 5 (5.2) |
|  | Multiple/Mixed | 4 (5.6) | 3 (4.9) | 2 (3.7) |
|  | Other ethnic group | 4 (9.1) | 1 (2.7) | 1 (3.2) |
| Role | Staff | 90 (6.4) | 68 (5.1) | 43 (3.6) |
|  | Post-graduate Research students | 25 (7.9) | 18 (6.8) | 11 (4.7) |
|  | Both | 8 (10.5) | 5 (7.5) | 3 (5.1) |

|  |  |  |  |
| --- | --- | --- | --- |
| Birthplace |  |  |  |
| United Kingdom | 76 (6.4) | 61 (5.9) | 36 (3.9) |
| EU | 28 (7.9) | 16 (5.2) | 13 (4.8) |
| Other | 16 (4.7) | 13 (4.2) | 7 (2.6) |
| Not provided | 3 (30.0) | 1 (9.1) | 1 (10.0) |
| Clinical role |  |  |  |
| Yes, COVID-19 contact<sup>1</sup> | 4 (7.4) | 4 (8.7) | 2 (5.9) |
| Yes, no COVID-19 contact | 7 (7.0) | 5 (5.9) | 4 (5.3) |
| No | 111 (6.4) | 82 (5.3) | 51 (3.7) |

<sup>1</sup>Part of the role contains contact with people with suspected Covid-19 symptoms.
\*A detailed list of participant self-reported demographics is available from the corresponding author.

## Discussion

This study found that over 90% of participants returned a valid test result across both periods of testing, with a reduction in the number of replacement kits required to be sent. This illustrates the feasibility of rapidly deploying low-cost SARS-CoV-2 serological testing without the need for face-to-face contact in an occupational setting. The initial findings described in this short report indicate that the positive antibody rate in our cohort was comparable to England population average at phase 1 [7]. Conversely, the positive antibody rate decreased over time, with the positive rate in phase 2 lower than the England population average [8]. However, it must be noted that the KCL CHECK study has not been designed as a representative cohort study.

Our study has some limitations. The SureScreen Diagnostics COVID-19 IgG/IgM Rapid Test Cassette used in this study was designed for “point-of-care” testing and, at the time of testing, rapid test cassettes had been certified by the Medicines and Healthcare products Regulatory Agency for use in laboratories using venous blood as there was insufficient data on their reliability when using capillary blood [9]. This may contribute to the level of invalid results observed and introduces additional uncertainty into the results compared with the laboratory-based validation. We relied on participants providing a clear photograph of the cassette result and recording the results within 10 minutes of applying the buffer solution, however, to overcome reporting issues, the research team developed a range of guidance documents to support participants in conducting and reporting the test results correctly which appear to have been effective.

In summary, we found that it was possible to conduct mass testing of postgraduate research students and members of staff at King’s College London using serological cassette testing on two separate occasions. We plan to repeat the antibody testing at regular intervals for two purposes (i) to continue to document changes in the antibody detection in our participants and to identify any associations with self-reported physical and mental health (ii) to look for patterns that can be used to reduce transmission risk within an occupational setting, and therefore add to the evidence about the potential utility of antibody testing for public health management purposes [10].

## Data Availability

Researchers may apply to have access to pseudonymised data. Requests to access study data is subject to submission of a research proposal to the Principal Investigators (Professor Matthew Hotopf, Professor Reza Razavi and Dr Sharon Stevelink). All requests must be made in accordance with the UK Policy Framework for Health and Social Care research. Where the applicant is outside of Kings College London, a data-sharing agreement is required.

## Acknowledgements

The research team would like to thank Jonathan Edgeworth for support during study setup. The research team would like to acknowledge Liam Jones, Lisa Sanderson, Jana Kim and Laila Danesh for providing support during packaging of the home testing kits. We would also like to thank Charlotte Williamson for providing support in annotating test result photographs. This paper represents independent research part funded by the National Institute for Health Research (NIHR) Biomedical Research Centre at South London and Maudsley NHS Foundation Trust and King’s College London. The views expressed are those of the author(s) and not necessarily those of the NHS, the NIHR or the Department of Health and Social Care.

